## Supplementary Materials for "Impact of Manganese on Neuronal Function: An Exploratory MultiOmic Study on Ferroalloy Workers in Brescia, Italy"

### Supplementary Data

**Table S1.** The set of metabolites with significantly biological (FC > 1.5) and statistical ($p < 0.05$) differences from volcano plot analysis in the student's t-test. Significantly upregulated metabolites (total=49) in the plasma of Mn exposed workers compared to control group, arranged in descending order from the highest to lowest fold change (FC). B. Significantly downregulated metabolites (total=13) in the plasma of Mn exposed workers compared to control group, arranged in descending order from the highest to lowest fold change (FC).

(a)

| Upregulated Metabolite | FC | p value |
|---|---|---|
| Oxymesterone | 5.844 | 0.025143 |
| HEPES | 5.561 | 0.04226 |
| diacetyl trimer | 5.031 | 0.024803 |
| Palmitoyl glutamic acid | 3.4201 | 0.013744 |
| Guanidinosuccinic acid | 3.174 | 0.016216 |
| LIC9QAS59U | 2.778 | 0.0053104 |
| 2-Hydroxy-2-methylbutyric acid | 2.774 | 0.031764 |
| Tetrahydrofuran | 2.774 | 0.031764 |
| C6 H6 N2 O2 | 2.7091 | 0.031643 |
| Butyric acid | 2.6534 | 0.0059909 |
| 2-Aminoisobutyric acid | 2.5645 | 0.014392 |
| (-)-pinellic acid | 2.5033 | 0.0089942 |
| C20 H37 N O4 S | 2.5013 | 0.046599 |
| C7 H6 O6 S | 2.4234 | 0.015619 |
| 1-O-palmitoyl-2-O-arachidonoyl-sn-glycero-3-phosphocholine | 2.157 | 0.015904 |
| L-(2,3-~2~H_2_)Threonine | 2.116 | 0.040442 |
| (S)-(+)-N-Boc-3-pyrrolidinol | 2.1056 | 0.044566 |
| 2-Deoxyhexopyranose | 1.9648 | 0.024223 |
| D-Mannoheptulose | 1.9648 | 0.024223 |
| (E)-Aconitic Acid | 1.9499 | 0.038327 |
| gamma-Glu-Ile | 1.9329 | 0.014855 |
| Purine | 1.8922 | 0.025603 |
| 2-Furoic acid | 1.8862 | 0.034909 |
| (±)5-iPF2?-VI | 1.8752 | 0.047639 |
| 2-Butoxyacetic acid | 1.8526 | 0.019614 |
| C5 H6 O4 | 1.843 | 0.026972 |

| Upregulated Metabolite | FC | p value |
|---|---|---|
| Choline | 1.8248 | 0.014724 |
| Pellitorin | 1.8167 | 0.018208 |
| 4-Octylphenol | 1.8167 | 0.018208 |
| ACETYL PROLINE | 1.7999 | 0.012912 |
| AMPA | 1.7595 | 0.0064805 |
| Ethylmalonic acid | 1.7443 | 0.0041374 |
| N-[(3S)-2-Oxotetrahydro-3-furanyl]pentanamide | 1.7305 | 0.0037919 |
| L-gamma-Glutamyl-L-valine | 1.7125 | 0.028958 |
| 2-(Methylamino)isobutyric acid | 1.6802 | 0.021439 |
| 2-Aminoethyl (2R)-3-[(1Z)-1-hexadecen-1-yloxy]-2-hydroxypropyl hydrogen phosphate | 1.6618 | 0.003523 |
| Hexacosanedioic acid | 1.6338 | 0.022585 |
| Trinonanoin | 1.6199 | 0.031432 |
| Homovanillic acid | 1.6157 | 0.035027 |
| (2S)-3-{[(1Z)-11-Methyl-1-dodecen-3-yn-1-yl]oxy}-1,2-propanediol | 1.583 | 0.015553 |
| PAF C-18-d4 | 1.5824 | 0.014784 |
| C20 H26 O | 1.5756 | 0.032239 |
| L(-)-Carnitine | 1.5749 | 0.015076 |
| DAz-1 | 1.5501 | 0.0061043 |
| Y-L-Glutamyl-L-glutamic acid | 1.5401 | 0.0046488 |
| Glu-Glu | 1.5356 | 0.0070337 |
| dilauroyl peroxide | 1.5213 | 0.041073 |
| C11 H16 N4 O5 | 1.5206 | 0.01287 |
| (5Z,8Z,11Z,14Z)-N-[2-(3,4-Dihydroxyphenyl)ethyl](5,6,8,9,11,12,14,15-~2~H_8_)-5,8,11,14-icosatetraenamide | 1.5188 | 0.0019601 |

(b)

| Downregulated Metabolite | FC | P value |
|---|---|---|
| 4-Allyl-2-hydroxy-6-methoxyphenyl hydrogen sulfate | 0.06805 | 0.031975 |
| C19 H37 N O4 | 0.11713 | 0.031942 |
| (2S,3aR,4R,5S,6R,6aS,7R,9aS,10aR)-9a-(Acetoxymethyl)-2,6,7-trihydroxy-7-isopropyl-4-methyl-1-methylenetetradecahydrodicyclopenta[ a, d][8]annulen-5-yl acetate | 0.24557 | 0.000174 |
| Sacubitril | 0.28154 | 0.004546 |
| C6 H12 O6 | 0.28408 | 0.013986 |
| C7 H8 O5 S | 0.29533 | 0.03716 |
| C6 H10 N3 O4 P | 0.36203 | 0.022048 |
| (2Z,6E)-N-(2-Hydroxyethyl)-2-methyl-2,6-icosadienamide | 0.48614 | 0.020388 |
| 16NS | 0.50839 | 0.025294 |
| 3,4-Dimethoxyphenethylamine | 0.5568 | 0.044327 |
| (3S)-3-{(Z)-[(3S)-3-{(Z)-[(3R)-3-Amino-1-hydroxy-4-methylpentylidene]amino}-1-hydroxybutylidene]amino}-5-methylhexanoic acid | 0.56918 | 0.044344 |
| Methyl 3-{(2S,3R)-3-[(2Z,5Z,8Z,11Z,14Z)-2,5,8,11,14-heptadecapentaen-1-yl]-2-oxiranyl}propanoate | 0.59735 | 0.024519 |
| N-[(2S,3R,4E,6E)-1-(beta-D-Glucopyranosyloxy)-3-hydroxy-4,6-pentadecadien-2-yl]octadecanamide | 0.60556 | 0.025875 |

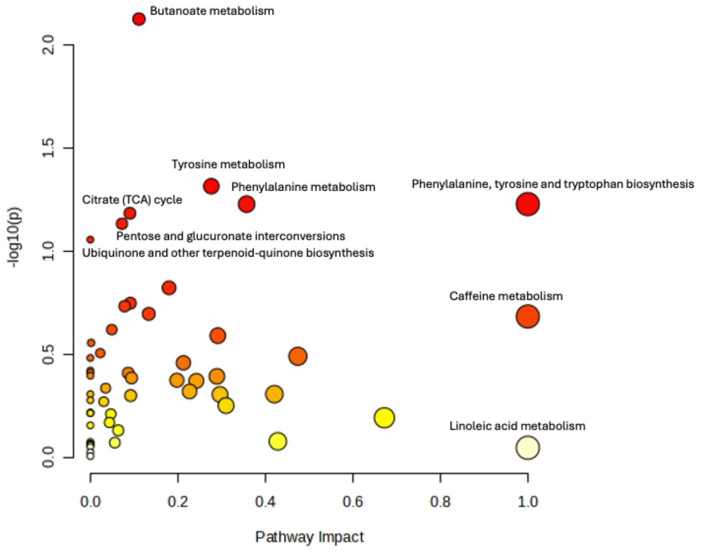


**Figure S1.** An overview of Pathway Analysis using distinct metabolites identified through a student's t-test was conducted by global test using Homo sapiens KEGG library. The X-axis indicating the impact or importance of respective pathways on Mn exposure study. The Y-axis describe the statistical significance of results (p-value). The size of bubble showing the number of compounds involved in the pathway that are affected by the exposure. Larger bubbles mean greater number of compounds. The bubble color represents the magnitude of p-value, a more intense red color indicates a lower p-value.

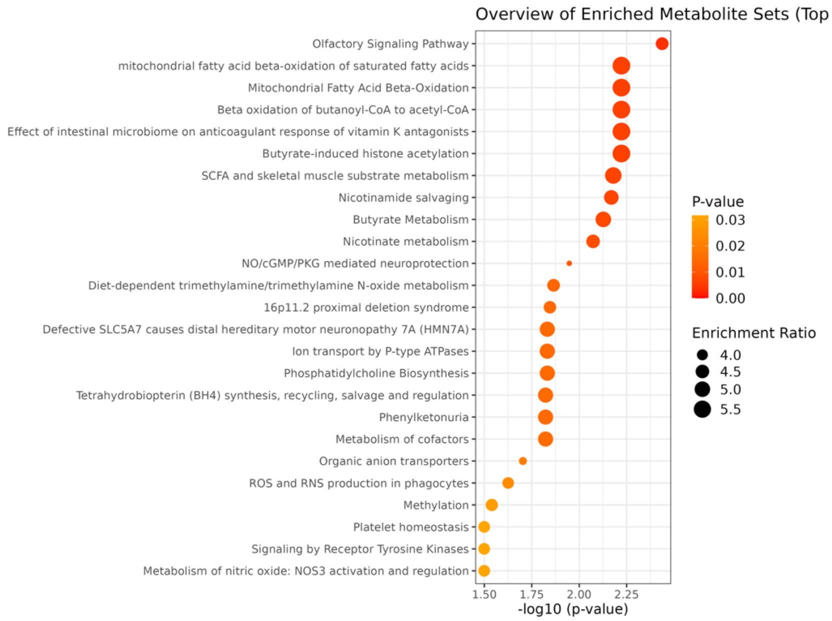


**Figure S2. Dot plot of Metabolomic Enrichment Analysis using RaMP-DB as a reference.** It indicates the top 25 enriched metabolite set in the study of metabolite profile of Mn-exposed workers than controls. The X-axis showing the p-value and Y-axis indicating metabolites set.

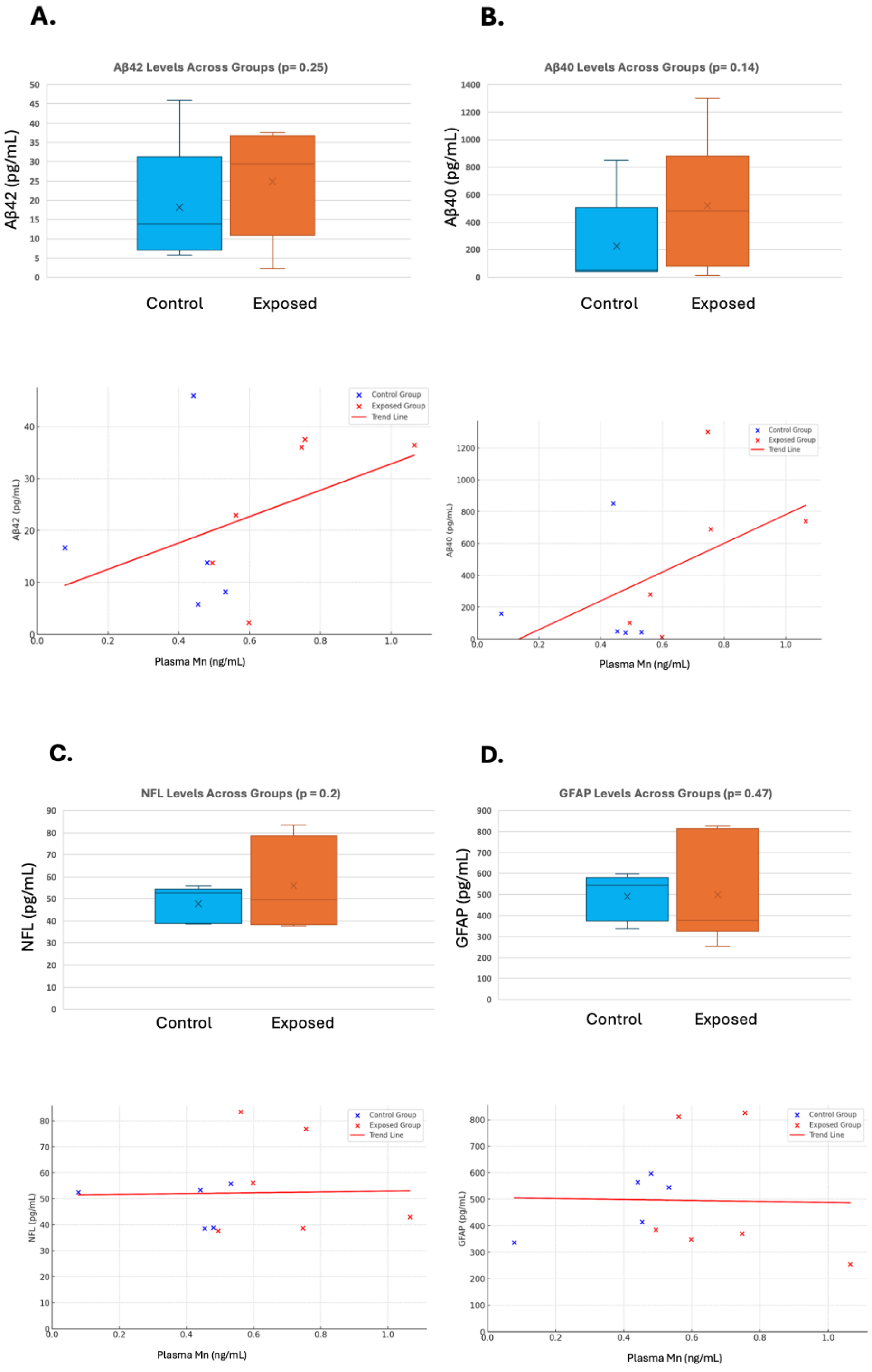


**Figure S3.** Comparative level of AD biomarker proteins. (a) Ab42 (b) Ab40 (c) NfL (d) GFAP Up**:** Bar chart of comparative level of proteins in plasma samples of Mn exposed workers and control group measured using single molecule arrays (Simoa) in Quanterix SR-X platform. Down: The Scatter plot represents the individual data points for proteins level versus PMn values. Blue points represent control individuals, while red points represent Mn exposed individuals. The red line indicating collective trend for both Mn exposed and control groups.

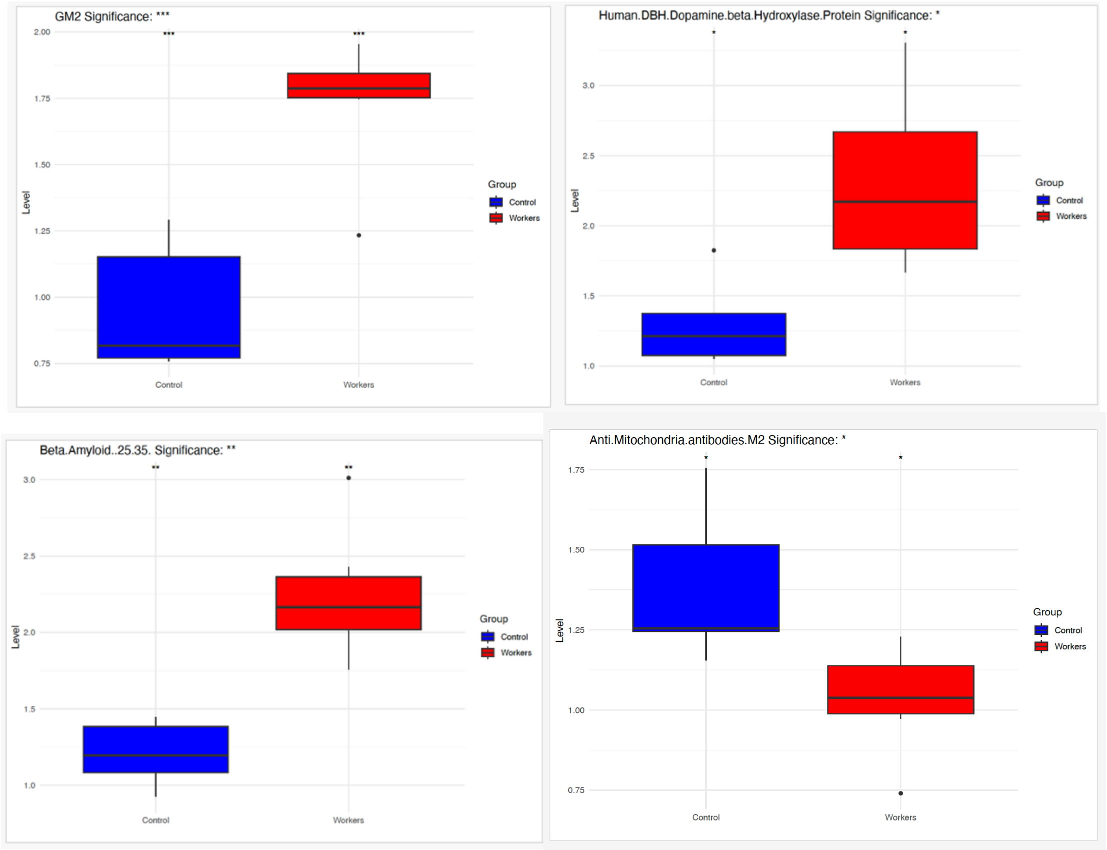


**Figure S4.** Bar plots of significantly distinct GM2 and dopamine beta-hydroxylase (DBH), Beta amyloid 25-35 and AMA-M2 antibodies. The antibodies pertinent to neurology exhibited significantly varied abundance levels ($p < 0.05$) in the plasma samples of the Mn-exposed group compared to controls, as assessed through protein chip analysis.

**Table S2. The panel of antigens employed in protein microarray analysis to assess plasma autoimmune antibody levels among Mn-exposed workers and control subjects.**

| AutoImmune Antigen | 3000/2675 | 3001/2849 | 3002 | 3003 | 3004/4755 | 3005/5608 | 3006/5965 | 3007/6013 | 3008/5997 | 3009/6069 | 3010/6166 |
|---|---|---|---|---|---|---|---|---|---|---|---|
| Group | Workers | Workers | Workers | Workers | Workers | Workers | Control | Control | Control | Control | Control |
| GM2 | 1.74613902 | 1.23335208 | 1.85631216 | 1.95385619 | 1.80451141 | 1.76957842 | 1.29186642 | 1.15181285 | 0.770845 | 0.81667917 | 0.758345 |
| Beta-Amyloid (25-35) | 1.75635799 | 2.42920375 | 2.16777418 | 3.01151218 | 1.97099894 | 2.16365473 | 1.08168179 | 1.44637487 | 1.38335458 | 1.19585167 | 0.92501417 |
| Tubulin Beta (TUBb) | 1.84831955 | 2.00003042 | 2.5 | 2.5290149 | 2.88130944 | 3.31325265 | 1.82843457 | 2.08081638 | 1.45835583 | 1.18335125 | 1.31668667 |
| 5-Hydroxytryptamine (5-HT) | 2.35808348 | 5.91259167 | 5.36544956 | 4.24372838 | 3.66272877 | 5.8659647 | 3.34073924 | 1.83345946 | 2.10419875 | 1.62919167 | 2.65420708 |
| Human DBH/Dopamine beta-Hydroxylase Protein | 1.79609397 | 1.66669208 | 1.94352179 | 2.39563807 | 2.76047347 | 3.30321264 | 1.37388963 | 1.82401852 | 1.21251875 | 1.07501667 | 1.05001625 |
| Human NOVA1 Protein | 1.8358311 | 1.8958625 | 2.41279078 | 2.36568673 | 2.39795913 | 3.04969994 | 1.85406815 | 1.36329323 | 1.51252333 | 1.80419417 | 1.67085875 |
| Human Siglec-1 / CD169 Protein | 2.15485887 | 3.02921292 | 4.33554836 | 3.45797434 | 2.93233175 | 3.58433787 | 2.93062237 | 1.91465197 | 1.88752875 | 1.95003 | 1.21251875 |
| Recombinant Adaptor Related Protein Complex 3 Beta 2 (AP3b2) | 2.02202486 | 1.98336375 | 1.92691034 | 1.85885438 | 2.55907644 | 2.16616473 | 1.38926892 | 2.16389858 | 1.27918625 | 0.86668 | 1.08335 |
| Anti Mitochondria antibodies M2 | 0.97269636 | 1.170018 | 1.04126677 | 1.22762151 | 0.74001137 | 1.03501587 | 1.51502319 | 1.255019 | 1.1550175 | 1.245019 | 1.75400625 |
| Human Brain Cerebellum Protein | 1.25681188 | 1.36252083 | 2.03073101 | 1.17512142 | 2.33351247 | 1.45080316 | 1.35167499 | 1.11216031 | 0.89168042 | 0.975015 | 0.88751375 |
| Recombinant Human GFAP Protein | 1.53496835 | 1.48335583 | 1.87707638 | 1.78491176 | 2.20730414 | 2.36194789 | 1.94292533 | 1.42749279 | 1.16251792 | 1.0416825 | 1.09168333 |
| Recombinant Human Amphiphysin/AMPH Protein | 1.96071677 | 2.67504083 | 5.0830577 | 2.01656676 | 2.72287737 | 3.45883527 | 1.23376648 | 2.33761386 | 1.39168792 | 1.98753042 | 1.30835333 |
| rhCarbonic Anhydrase VIII | 3.24477733 | 2.7750425 | 4.65116231 | 2.98109286 | 3.61976488 | 3.81275055 | 3.09637859 | 2.65105701 | 2.77920917 | 2.74170875 | 1.10835042 |
| Recombinant Leucine-rich glioma-inactivated protein 1 (LGI1) | 2.70322347 | 2.1375325 | 1.76079743 | 1.18214163 | 1.78571444 | 1.75702816 | 1.28503109 | 2.03549795 | 1.00834875 | 0.88751375 | 0.84167958 |
| Recombinant Human RAGE Protein | 2.10830986 | 1.42918833 | 1.71511623 | 1.33470621 | 1.11439305 | 1.566265 | 0.73479175 | 1.73904869 | 0.87084667 | 0.98334833 | 0.95001458 |
| PDC-E2 | 1.2627986 | 1.190018 | 0.74616138 | 0.97186704 | 0.76501175 | 1.19501831 | 1.33002037 | 1.1950185 | 1.040016 | 1.31002 | 1.72369012 |
| PM/Scl75 | 0.88737215 | 1.045016 | 2.42802203 | 1.35976132 | 0.78501205 | 1.11001701 | 1.67502564 | 1.8000275 | 1.2600195 | 2.9000445 | 2.6808146 |
| Human STK23/MSSK1/SRPK3/SPRY3 recombinant protein | 2.36262474 | 2.26253458 | 3.90780739 | 2.73633435 | 1.57626221 | 1.89759052 | 2.08304967 | 1.5143508 | 1.97503 | 1.95836333 | 1.2458525 |
| Ribosomal P1 | 1.11774748 | 1.100017 | 1.07005759 | 1.4620631 | 0.76001167 | 1.13001732 | 1.49002281 | 1.4650225 | 1.240019 | 1.845028 | 1.02208754 |
| U1-snRNP C | 4.54778056 | 1.6500255 | 6.64347364 | 6.25319679 | 0.85001305 | 2.10003212 | 2.07003166 | 1.9200295 | 1.0750165 | 1.910029 | 1.50714599 |
| Human GRM1 / MGLUR1 Protein | 1.65077151 | 1.71252625 | 1.47425239 | 1.78023194 | 3.40494001 | 2.67319214 | 1.81134665 | 2.03738614 | 1.33335375 | 0.92918083 | 1.14168417 |
| Recombinant Bone Morphogenetic Protein 1 (BMP1) | 3.09037236 | 1.9208625 | 1.85631216 | 0.52087229 | 3.47475903 | 2.02058237 | 1.48325401 | 1.46336876 | 1.47085583 | 0.92501417 | 1.29168625 |
| Ribosomal P2 | 1.14334479 | 1.0800165 | 1.15643008 | 1.08695655 | 0.7550116 | 1.06501633 | 1.53002342 | 1.5350235 | 0.9950155 | 2.3900365 | 1.16067573 |
| POLY(L-GLUTAMIC ACID) | 1.33742031 | 1.65835875 | 1.59883728 | 1.02957705 | 1.726638 | 2.24146579 | 1.42686303 | 1.16125398 | 1.17918458 | 0.88751375 | 1.60835792 |
| Recombinant Human NARG2 Protein (NMDA) | 5.26907282 | 4.2959 | 5.71843836 | 2.98717663 | 4.31525373 | 5.10793159 | 2.15652772 | 6.02152724 | 3.66255583 | 1.78336042 | 1.80419417 |
| U1-snRNP A | 3.37030746 | 2.7250415 | 8.64203484 | 5.2344415 | 0.74501144 | 1.19501831 | 2.35503601 | 1.565024 | 1.31002 | 1.820028 | 0.9874406 |
| Anti angiotensin II type1 receptor | 1.62969276 | 1.1500175 | 1.51871415 | 1.34271102 | 0.80001228 | 1.0700164 | 1.84502823 | 1.9950305 | 1.105017 | 1.2200185 | 2.70246866 |
| Histidyl-tRNA Synthetase(Jo-1) | 21.8088743 | 6.080095 | 22.1473137 | 13.4782605 | 1.20001839 | 2.36003609 | 8.48012947 | 4.985076 | 1.4800225 | 4.115063 | 4.20528384 |
| Recombinant Human GAD2/GAD65 Protein | 3.12897303 | 3.14171458 | 1.8978406 | 3.25346279 | 4.59452176 | 5.01254901 | 2.5 | 4.78096819 | 1.30418667 | 0.97918167 | 1.61252458 |
| Recombinant Human Enolase 2/Neuron-specific Enolase Ptotein | 4.02247876 | 4.4709 | 6.96428726 | 4.05934053 | 3.44253611 | 3.05973995 | 2.8827762 | 3.20052945 | 4.6667375 | 2.01253083 | 3.57922125 |
| Recombinant Human Dopamine D2R/DRD2 | 2.51930034 | 3.75422417 | 3.4260797 | 3.90256411 | 2.98066614 | 2.07831316 | 1.33629552 | 4.24848945 | 2.25003458 | 1.51252333 | 1.30418667 |
| Human ELAVL3 / HUC Protein | 3.48319667 | 2.46670417 | 3.77491698 | 3.62457839 | 2.05692808 | 1.61646579 | 2.05912488 | 3.05136063 | 2.52087167 | 1.61252458 | 1.44168875 |
| Formiminotransferase Cyclodeaminase (LC1) | 7.12884194 | 3.1100475 | 7.54078723 | 4.47570324 | 1.14501755 | 1.73502655 | 3.71005669 | 3.150048 | 1.4600225 | 1.250019 | 2.37332209 |
| Recombinant GRB2 Related Adaptor Protein 2 (GRAP2) | 3.31743856 | 2.67087417 | 1.83139518 | 1.95993996 | 2.93233175 | 9.22439738 | 1.35680137 | 3.73489449 | 1.74586 | 1.23751875 | 1.07918333 |
| U1-snRNP B/B’ | 2.72184336 | 1.2800195 | 6.12763937 | 5.75447558 | 0.68501053 | 1.29501984 | 2.2100338 | 1.690026 | 0.860013 | 1.2650195 | 1.2906023 |
| Recombinant Human NRXN3 Protein | 2.5 | 3.32088417 | 4.70930095 | 4.18008185 | 2.75778823 | 2.93674735 | 2.90840808 | 3.57439653 | 1.96253 | 3.27921667 | 1.408355 |
| Human Tau-441 / 2N4R Protein | 6.41008041 | 5.02090833 | 4.99999834 | 4.70656853 | 2.68528396 | 4.6234937 | 3.7491456 | 4.37311181 | 4.62090417 | 4.2334 | 1.56669083 |
| CENPB | 1.20733784 | 1.895029 | 2.68713996 | 1.39386191 | 0.86501328 | 1.38502121 | 2.19003349 | 1.96503 | 1.255019 | 2.5050385 | 4.96319004 |
| Fibulin | 1.75767929 | 1.715026 | 1.40595027 | 2.42540493 | 1.20001839 | 2.49503815 | 4.70507187 | 2.3300355 | 1.4050215 | 2.7750425 | 2.27804246 |
| Recombinant Human Titin protein | 2.54541256 | 2.27503458 | 4.52242673 | 2.8818792 | 2.53759395 | 10.4216858 | 3.18352697 | 2.39614772 | 1.93752958 | 1.35002083 | 2.52920542 |
| Recombinant Human AIF-1/Iba1 Protein | 3.30267834 | 3.95006042 | 5.20764051 | 3.87588865 | 2.65843179 | 3.36596269 | 4.01913984 | 2.65483339 | 3.55005417 | 3.06671333 | 1.27501958 |
| Recombinant Calcium Channel Alpha 1B Subunit (CACNa1B) | 4.93528483 | 25.7003917 | 23.1021594 | 4.81233721 | 4.70461944 | 6.90010052 | 4.90430766 | 6.06117921 | 10.404325 | 3.25838292 | 3.57922125 |
| Recombinant Human MBP Protein | 3.54677539 | 3.29588375 | 5.2200988 | 3.23895534 | 1.37218047 | 3.29568264 | 3.40909091 | 3.24962236 | 1.96669667 | 2.52087167 | 1.08335 |
| Nucleolin | 10.9556309 | 4.000061 | 4.42178484 | 36.2787713 | 1.38002113 | 2.39503662 | 3.84005867 | 3.080047 | 1.575024 | 2.0550315 | 1.99653526 |
| Proliferating Cell Nuclear Antigen | 1.11774748 | 1.0800165 | 2.05374294 | 1.15089517 | 0.84001289 | 1.19001823 | 1.62502487 | 1.5450235 | 0.985015 | 2.0650315 | 1.49848437 |
| SmD2 | 5.07252513 | 4.4250675 | 11.1708255 | 17.7195221 | 1.03501587 | 2.22003395 | 6.32009651 | 4.435068 | 1.5850245 | 1.900029 | 2.8757038 |
| Nup62 | 2.35068263 | 1.6600255 | 1.30518237 | 1.50042627 | 0.81001244 | 1.32502029 | 6.08009285 | 1.570024 | 1.29002 | 1.450022 | 3.13555693 |
| TIF1 gamma | 0.93856676 | 0.8900135 | 2.1353168 | 1.17220804 | 0.78001198 | 1.06501633 | 1.44502213 | 1.6550255 | 0.835013 | 1.7200265 | 5.04114468 |
| HLA–B*14:02 | 2.242535 | 2.33949522 | 1.91335713 | 1.71365543 | 2.0179372 | 2.00607155 | 1.81306453 | 1.93687583 | 1.85334155 | 2.00328195 | 2.0062124 |
| Cluster of differentiation 11b (CD11b) | 2.46593945 | 1.86252833 | 1.66528227 | 1.54483308 | 2.05692808 | 2.86646479 | 2.22146352 | 2.61706961 | 1.25418583 | 0.93751417 | 1.10001667 |
| GP2 | 2.32935147 | 1.575024 | 7.34165158 | 2.67263425 | 7.15010918 | 1.52002327 | 2.26003456 | 2.105032 | 1.0750165 | 2.365036 | 4.03637952 |
| Recombinant Human CD68/SR-D1 Protein | 2.75544905 | 1.75419333 | 2.75747489 | 2.98951654 | 1.16272824 | 1.29769079 | 0.87662352 | 2.41691968 | 2.44170417 | 1.37085417 | 0.65834333 |
| U1-snRNP 68/70 kDa | 4.45819149 | 5.21508 | 9.73848407 | 6.31287284 | 0.93001427 | 3.71505676 | 7.73011803 | 2.095032 | 2.2450345 | 2.4450375 | 1.63274175 |
| Recombinant Human Recoverin Protein | 3.96003536 | 3.56255458 | 5.75996596 | 3.37981989 | 2.07572505 | 2.14859447 | 2.76828544 | 3.63481858 | 3.47088625 | 2.64587375 | 0.92918083 |
| Recombinant Human S100B | 3.73410447 | 66.226 | 4.64285679 | 6.88459106 | 2.8974209 | 2.85391478 | 3.46377396 | 3.06835433 | 3.48338667 | 2.97921208 | 1.63335833 |
| Proteinase 3 | 3.32764514 | 1.170018 | 3.01583457 | 1.4791134 | 0.80501236 | 1.20001839 | 1.37502106 | 1.2000185 | 1.0150155 | 1.6100245 | 1.55911662 |
| RNA polymerase III | 3.22525634 | 1.710026 | 4.89923198 | 2.78346119 | 0.90001381 | 1.10501694 | 4.98507614 | 2.0050305 | 1.165018 | 3.675056 | 21.9055873 |
| BPI | 3.70307201 | 1.2850195 | 4.89443359 | 13.2011932 | 0.79001213 | 1.09001671 | 1.58002419 | 1.055016 | 0.9300145 | 1.8550285 | 4.66868607 |
| DFS70 | 11.3865203 | 11.05517 | 7.34884917 | 4.03665808 | 0.9650148 | 3.63505554 | 5.14507858 | 2.8450435 | 2.6450405 | 5.82009 | 5.99826741 |
| Gp210 | 2.78157026 | 1.120017 | 1.0892514 | 1.21483379 | 0.78001198 | 1.08001656 | 3.00504593 | 1.645025 | 1.1300175 | 1.180018 | 2.01818976 |
| Anti Mitochondria antibodies M4 | 2.03071694 | 1.630025 | 1.41074866 | 1.79454391 | 0.81501251 | 1.52502335 | 1.79502747 | 1.640025 | 1.31002 | 1.570024 | 3.94110033 |
| HLA–C*08:02 | 22.3942924 | 15.722608 | 12.8028421 | 2.72463095 | 3.32735478 | 9.83485254 | 14.18481 | 42.8294895 | 12.903613 | 20.2486708 | 2.39082733 |
| glucose transporter 2 | 2.97781599 | 1.230019 | 9.91602695 | 4.4501278 | 0.78501205 | 1.4300219 | 3.32505081 | 2.8350435 | 0.9300145 | 1.175018 | 1.73668256 |
| HLA–C*17:01 | 13.0620432 | 3.37218864 | 5.74207696 | 2.55837718 | 2.53811633 | 3.77170595 | 2.82527975 | 3.97114999 | 3.01005378 | 4.63014446 | 3.04768819 |
| Sm/RNP | 1.46757705 | 0.910014 | 9.19385705 | 4.0323955 | 0.67501038 | 1.19501831 | 2.07503174 | 1.6700255 | 1.100017 | 1.7450265 | 1.22996961 |
| Human KRT6A / CK6A / Cytokeratin 6A Protein | 2.03006984 | 3.34304693 | 2.8289207 | 2.2296348 | 2.31133952 | 3.31876201 | 2.6702072 | 2.43743037 | 4.99704321 | 3.9308266 | 2.17430957 |
| Recombinant Human CRMP5 Protein | 3.14032617 | 4.6917375 | 4.92109589 | 3.21789566 | 1.44468312 | 13.792671 | 9.55229142 | 4.89426142 | 4.18339583 | 27.6045875 | 3.02921292 |
| HA (Tyrosyl tRNA Synthetase) | 42.7602276 | 5.97009 | 7.31286124 | 21.4791128 | 1.19001823 | 3.84505875 | 4.88507462 | 10.050155 | 1.3300205 | 4.2100645 | 16.2884358 |
| AQP4 Recombinant | 4.13601243 | 11.1001708 | 6.80232545 | 4.07899626 | 1.93340499 | 12.2138548 | 9.47026941 | 6.28587748 | 3.48338667 | 2.87087708 | 1.59169083 |
| Platelet Glycoprotein | 11.2073378 | 3.135048 | 0.69817674 | 11.4961634 | 1.72002632 | 3.17504852 | 7.04510758 | 3.480053 | 2.035031 | 2.4650375 | 1.30792555 |
| ß2-Glycoprotein 1 (Apolipoprotein H) | 25.6868613 | 5.515085 | 12.3392506 | 15.3282178 | 1.21001854 | 2.81504303 | 9.42514389 | 11.74518 | 1.575024 | 3.065047 | 7.80424689 |
| Recombinant Human Galectin 3 Protein | 3.46843759 | 3.83339208 | 5.0041511 | 2.94084591 | 2.51611173 | 4.7540163 | 2.75974148 | 4.06155685 | 4.89590833 | 2.28336833 | 1.6791925 |
| Recombinant Human CDR2 Protein | 2.73274277 | 22.1253375 | 4.90863761 | 2.3947021 | 3.86949522 | 5.66766953 | 7.50854567 | 3.17975937 | 3.57088792 | 3.57922125 | 2.92921125 |
| Dopamine Receptor D1 (DRD1) Protein | 7.48751041 | 2.45420417 | 4.44767289 | 2.5 | 1.93340499 | 5.18323415 | 59.9367952 | 2.5 | 3.77089083 | 2.89171083 | 1.82919458 |
| Collagen Complex - 1 | 3.12713326 | 4.7500725 | 1.42514407 | 2.86018753 | 0.93501434 | 1.69502594 | 3.22004921 | 2.040031 | 1.2700195 | 1.505023 | 1.52013887 |
| Cytochrome P450 2D6 (LKM 1 hp) | 8.52815936 | 2.415037 | 2.43042123 | 1.43648766 | 0.84501297 | 1.22501877 | 2.18003334 | 2.750042 | 1.235019 | 1.225019 | 1.57210949 |
| ASCA | 3.11433482 | 1.4050215 | 6.45393478 | 14.2199485 | 0.83001274 | 1.21001854 | 1.93002953 | 2.940045 | 1.1300175 | 1.175018 | 6.01125985 |
| TRP1 Protein | 24.2150194 | 2.305035 | 1.85460658 | 6.94799643 | 0.64000984 | 1.14501755 | 2.38503647 | 9.525145 | 0.8150125 | 1.3400205 | 0.51537458 |
| alpha-2B Adrenergic R/ADRA2B Recombinant Protein Antigen | 1.46571306 | 1.204185 | 1.75664467 | 1.87710569 | 1.36143949 | 1.18975895 | 1.39952185 | 2.12424472 | 1.12918417 | 0.90834708 | 0.76251167 |
| SS-A 60kDa | 2.24829383 | 1.2850195 | 4.3905953 | 4.29241254 | 1.01501556 | 2.1250325 | 2.20003365 | 2.430037 | 1.245019 | 2.62004 | 1.77566075 |
| FHL1 Protein | 20.5716747 | 24.055365 | 1.44913627 | 7.52344398 | 0.73001122 | 5.04507706 | 5.4150827 | 20.035305 | 1.30002 | 1.2700195 | 0.82286709 |
| Recombinant Endothelin Receptor A (ETRA) | 4.33015339 | 47.279875 | 10.1287364 | 6.21115513 | 3.27604831 | 7.40712843 | 6.02529217 | 13.549852 | 5.95009167 | 5.09174583 | 7.67511667 |
| Caspr2 Recombinant Protein Antigen | 5.1271563 | 2.35003583 | 1.51162806 | 2.47519647 | 2.78195542 | 2.64558212 | 33.8294639 | 2.44335433 | 1.07084958 | 0.96668167 | 1.2583525 |

| | | | | | | | | | | | |
|---|---|---|---|---|---|---|---|---|---|---|---|
| Parietal Cell (H/K-ATPase-α, βsubunits) (PCA) | 8.35750923 | 1.705026 | 1.39635325 | 1.53026429 | 1.12001717 | 1.11501709 | 5.74008766 | 1.435022 | 1.1950185 | 4.330066 | 5.68644452 |
| Ku(p70/p80) | 1.03668942 | 0.855013 | 3.18857906 | 1.35123617 | 0.69501068 | 1.08501663 | 2.37503632 | 1.380021 | 0.9300145 | 2.1300325 | 1.71935931 |
| SAE1/SAE2 Human Recombinant | 27.9820824 | 2.230034 | 1.6026872 | 4.96163673 | 0.8800135 | 1.240019 | 4.45006798 | 7.71512 | 0.905014 | 1.4050215 | 0.73624953 |
| Human Keratin 18 (KRT 18) Protein | 1.7921932 | 1.82132811 | 1.6225433 | 1.89067036 | 1.92825119 | 1.79235045 | 1.64099915 | 1.89618973 | 2.03666461 | 1.88967473 | 1.86278049 |
| HLA–C*07:02 | 25.3074051 | 12.0553661 | 23.8718359 | 2.87205362 | 4.26137129 | 10.9908933 | 12.8900764 | 44.0315547 | 11.2383219 | 19.9886394 | 6.42700382 |
| INS-IGF2 | 21.3907877 | 6.965105 | 3.57245757 | 25.3239549 | 1.7250264 | 5.03507691 | 15.8002412 | 9.69015 | 3.1750485 | 5.29008 | 2.34300596 |
| Human KRT10 / CK10 / Cytokeratin 10 Protein | 10.9726955 | 1.640025 | 0.7797506 | 1.33418587 | 0.82501266 | 1.23501892 | 1.96503006 | 2.885044 | 1.170018 | 1.095017 | 0.81420547 |
| HLA–A*33:01 | 1.31643921 | 1.93516027 | 1.40393053 | 1.11481767 | 1.66431799 | 1.74620543 | 1.33616572 | 1.60399471 | 1.68775919 | 1.60944166 | 1.97515039 |
| Sm | 1.95819117 | 1.120017 | 6.13243776 | 10.899403 | 0.78001198 | 1.59002434 | 3.88505936 | 1.770027 | 0.990015 | 2.560039 | 3.43005656 |
| Tyrosinase Recombinant Protein Antigen | 31.6040963 | 5.510085 | 1.07725542 | 25.0682004 | 2.26003456 | 11.2051711 | 124.716903 | 7.510115 | 3.530054 | 8.830135 | 6.76050355 |
| Human MAG/Siglec-4a Protein | 4.03950847 | 3.32505083 | 15.7931889 | 2.89872703 | 2.22073036 | 1.24246999 | 1.92241983 | 4.94146614 | 2.70420792 | 5.20424583 | 1.65002542 |
| Muscarinic M2 Acetylcholine receptor | 1.90699655 | 1.2750195 | 0.95969291 | 1.44501281 | 1.33502045 | 1.1350174 | 1.29501984 | 1.250019 | 0.985015 | 1.575024 | 2.8237336 |
| Ribosomal P0 | 1.2627986 | 1.29002 | 1.02927079 | 2.22080136 | 0.76001167 | 1.32002022 | 2.04503128 | 1.780027 | 0.8650135 | 1.705026 | 1.13902123 |
| HLA–B*07:02 | 1.87972063 | 2.39140317 | 2.02166072 | 2.02733241 | 2.08584264 | 2.23315129 | 2.14445093 | 2.01578104 | 2.07096451 | 1.83539507 | 3.46976006 |
| Beta1 Adrenergic Receptor | 8.08020543 | 2.0050305 | 0.50623797 | 1.21483379 | 0.78001198 | 1.10001686 | 1.50002296 | 2.435037 | 1.30502 | 1.685026 | 0.77522728 |
| Glycine Receptor Alpha 1 Recombinant Protein | 7.81221475 | 2.8833775 | 1.56561478 | 1.3992887 | 4.45757165 | 3.76757051 | 28.482569 | 2.43391338 | 0.95418125 | 0.89168042 | 2.41670375 |
| HMGCR | 3.01194559 | 1.0700165 | 1.58589259 | 2.30179027 | 0.77001183 | 1.02501572 | 1.17001793 | 1.430022 | 1.0700165 | 1.97503 | 4.88090025 |
| Recombinant Human PNMA1 Protein | 2.98478588 | 5.970925 | 7.2799012 | 4.08086819 | 2.92159078 | 8.17520154 | 8.05878517 | 4.09365606 | 3.03337958 | 2.54587208 | 4.55840417 |
| Recombinant Human PNMA2 Protein | 2.39554884 | 2.44170417 | 2.44186052 | 1.71284155 | 2.69065445 | 1.94779131 | 2.99555817 | 3.17787118 | 1.43335542 | 1.14168417 | 1.204185 |
| Recombinant Human BACE-1 Protein | 3.58310544 | 4.045895 | 4.53903778 | 3.40228351 | 2.59667038 | 3.6244979 | 6.09022626 | 2.91163275 | 7.004275 | 3.44588583 | 1.73752667 |
| Fibrillarin | 2.68344718 | 1.190018 | 3.36852114 | 16.9437335 | 0.89001366 | 1.31502014 | 1.72002632 | 3.3800515 | 1.7200265 | 2.540039 | 5.03248305 |
| GAD II (GAD 65 kD) | 1.24573401 | 1.385021 | 2.06573916 | 1.53026429 | 0.83001274 | 1.30001991 | 1.78502731 | 1.2200185 | 1.1400175 | 2.025031 | 1.45084455 |
| Collagen Complex - 3 | 3.70733816 | 2.7750425 | 0.9237045 | 1.10400685 | 0.68501053 | 1.10001686 | 2.03503113 | 1.9350295 | 0.8100125 | 1.2050185 | 1.03941122 |
| Tropomyosin | 35.4522203 | 15.80024 | 3.45249539 | 28.2097178 | 1.01001549 | 6.30509628 | 24.1903692 | 15.77024 | 1.8500285 | 4.58007 | 9.56258309 |
| ZO (Phenylalanyl tRNA Synthetase) | 1.42918088 | 1.0900165 | 1.94097907 | 1.83716966 | 0.8600132 | 1.34002052 | 1.21001854 | 1.620025 | 1.055016 | 1.560024 | 2.42096147 |
| KRT14 Human | 35.4180911 | 3.3600515 | 1.06046081 | 11.2745095 | 0.97501495 | 2.7800425 | 9.48514481 | 15.515235 | 1.2200185 | 2.030031 | 0.85751403 |
| SmD | 1.31825936 | 0.9400145 | 12.555183 | 1.99062234 | 0.74501144 | 1.23001884 | 2.52003853 | 1.830028 | 1.105017 | 1.890029 | 3.12689487 |
| SmD1 | 0.94283291 | 0.775012 | 16.6242813 | 1.61977836 | 0.60000923 | 0.82501266 | 1.89002892 | 1.0850165 | 0.925014 | 1.6050245 | 5.17539989 |
| Recombinant Human SNCA | 1.62465929 | 1.00834875 | 1.66943504 | 1.07918364 | 1.32921603 | 0.97389553 | 3.01264608 | 1.31797613 | 1.43335542 | 0.80834583 | 0.86251333 |
| Recombinant Human Mannan Binding Lectin/MBL Protein | 4.1984547 | 3.28755042 | 5.46926858 | 3.88197289 | 3.2626221 | 3.12751 | 3.51162013 | 4.32590709 | 2.78337583 | 5.34591667 | 1.65835875 |
| 21 Hydroxylase | 5.08532357 | 2.095032 | 0.85892525 | 5.78857618 | 1.22001869 | 3.75505737 | 10.3451579 | 2.1900335 | 2.1350325 | 3.27505 | 2.01385895 |
| Human KCNA1 Protein | 3.34468609 | 5.30424583 | 6.88538066 | 4.35136619 | 3.49892622 | 7.55271104 | 5.59296101 | 7.33950205 | 5.7500875 | 2.41253708 | 1.74586 |
| Human PLP1 / Myelin PLP Protein Recombinant | 3.80335976 | 6.57926667 | 9.05315484 | 3.58667136 | 5.91836838 | 6.48343268 | 4.71633714 | 9.61480567 | 4.62507083 | 12.4085208 | 2.733375 |
| Recombinant Human Neurofascin Protein | 2.36603068 | 3.12088083 | 3.97009963 | 3.08451883 | 3.12298674 | 2.30421684 | 3.20403418 | 4.32968346 | 2.60837333 | 3.46671958 | 2.33336917 |
| CENPA | 1.42918088 | 1.9150295 | 6.57629616 | 1.20630864 | 1.09501678 | 1.64502518 | 4.40006722 | 2.215034 | 1.1300175 | 3.135048 | 2.91468198 |
| Recombinant Human GRP78/HSPA5 Protein | 3.16303359 | 3.07504708 | 4.71760648 | 1.9412202 | 2.95918419 | 3.17520004 | 4.10970751 | 2.95128472 | 3.63338875 | 2.20003375 | 1.73336 |
| Glutamate Decarboxylase -GAD I (GAD 67 kD) | 1.70221853 | 1.3450205 | 3.99472081 | 3.78942876 | 0.81501251 | 1.29501984 | 2.07503174 | 1.5350235 | 1.4000215 | 2.740042 | 4.84625374 |
| F- actin | 1.36092168 | 1.180018 | 4.74088267 | 1.24893438 | 1.08001656 | 1.16501785 | 1.30001991 | 1.4000215 | 1.1450175 | 1.94503 | 1.92724138 |
| Asparaginyl-tRNA Synthetase (KS) | 0.94283291 | 1.97003 | 3.70921412 | 6.00596747 | 0.95001457 | 1.39502136 | 2.22503403 | 1.0850165 | 0.990015 | 3.0300465 | 3.26115224 |
| Oncomodulin Recombinant Protein | 201.63396 | 13.75521 | 0.59500965 | 13.934356 | 1.90002907 | 4.53506928 | 101.756553 | 16.22525 | 1.710026 | 2.3750365 | 0.97011691 |
| Nuclear Matrix Protein 2 (NXP2) | 1.85153579 | 0.990015 | 8.99712061 | 3.0306905 | 0.69001061 | 1.10501694 | 1.67002556 | 2.2400345 | 1.035016 | 2.1850335 | 4.11000465 |
| Human SEPT5 / Septin 5 Protein | 13.0177032 | 33.217175 | 16.6694338 | 11.8831899 | 3.03168577 | 1.16967869 | 5.6151753 | 31.1480378 | 37.725575 | 3.67505625 | 4.20839583 |
| Human PMEL/SILV/gp100 protein | 3.28015799 | 5.26454745 | 6.65162378 | 3.67427037 | 5.27994897 | 5.67091696 | 2.70419607 | 3.79607931 | 3.40508624 | 13.8197477 | 5.082221 |
| SS-B (La) | 2.43600684 | 1.4000215 | 3.79798436 | 1.913896 | 0.8800135 | 1.31002007 | 3.40505203 | 1.555024 | 1.2600195 | 2.5300385 | 2.01385895 |
| Human Melan-A/MART-1 Protein | 29.6587036 | 10.45516 | 8.97792704 | 8.45268521 | 1.04501602 | 4.79507324 | 8.95513672 | 32.16549 | 3.1700485 | 1.845028 | 18.5361626 |
| Glycyl-tRNA Synthetase (EJ) | 2.77730369 | 3.220049 | 2.24328228 | 2.23785165 | 0.88501358 | 1.23501892 | 1.84502823 | 1.500023 | 1.1450175 | 2.155033 | 3.0619318 |
| Mi-2 | 0.83617754 | 1.2100185 | 12.4304224 | 1.51747657 | 0.95501465 | 1.09001671 | 1.77502716 | 1.7300265 | 0.9600145 | 2.3350355 | 5.0151598 |
| Human EEF1A1 Recombinant Protein | 1.53269772 | 1.82086125 | 1.43272437 | 1.57759231 | 1.58700318 | 2.10090368 | 2.29323277 | 2.44524252 | 2.4625375 | 0.82084583 | 0.975015 |
| SUMO | 44.8336175 | 11.03517 | 2.80470295 | 18.1457795 | 1.05501617 | 3.76005745 | 8.68013252 | 38.900595 | 2.285035 | 1.8050275 | 0.92247753 |
| Scl-70 | 1.26706475 | 1.1950185 | 4.35220817 | 1.78601876 | 0.82501266 | 1.31002007 | 1.78502731 | 1.8100275 | 0.9400145 | 2.61504 | 2.76743216 |
| Recombinant Human Beta-amyloid 42 | 4.64463991 | 4.46673333 | 4.90033209 | 4.6710028 | 2.33619772 | 1.40311237 | 2.98530542 | 4.66389858 | 5.2209125 | 2.52920542 | 1.94169625 |
| SS-A 52kDa (Ro-52) | 2.06484654 | 2.290035 | 5.96929006 | 3.52088657 | 1.26501938 | 2.12003243 | 3.31005058 | 2.415037 | 2.2500345 | 3.0500465 | 2.24339552 |
| SLA (Soluble Liver Antigen) | 2.35921535 | 1.115017 | 1.71305188 | 3.06052852 | 0.9650148 | 1.02001564 | 1.48002266 | 1.2850195 | 1.165018 | 1.7400265 | 3.72022539 |
| BCOADC-E2 | 1.99232077 | 1.4100215 | 1.55950095 | 5.68201182 | 0.87001335 | 1.21501862 | 3.26004982 | 1.4850225 | 1.3950215 | 1.375021 | 1.90558688 |
| Recombinant Human HOMER3 protein | 4.16439528 | 7.8167875 | 6.79401993 | 3.87495269 | 2.55370568 | 1.31777105 | 3.25529794 | 3.14010551 | 8.7168 | 2.87504375 | 1.95419667 |
| Islet Cell Antigen (IA-2) | 3.61348123 | 1.7350265 | 0.5542226 | 1.41091221 | 0.82001259 | 1.38502121 | 1.81502777 | 2.2050335 | 1.045016 | 1.2650195 | 0.95712447 |
| HLA–B*42:01 | 2.26582892 | 2.01620993 | 2.22121943 | 2.16237983 | 1.69250462 | 1.86520909 | 2.08709545 | 1.89865671 | 1.82968726 | 2.38197477 | 1.81070656 |
| Alanyl-tRNA Synthetase (PL-12) | 1.72354969 | 1.8500285 | 1.41074866 | 3.2821824 | 1.57002403 | 1.48502274 | 1.88002876 | 1.4750225 | 1.4100215 | 1.6500255 | 3.63793864 |
| Recombinant Human TRP2 Protein | 27.9266224 | 11.15517 | 0.68378133 | 4.19011076 | 1.77002708 | 10.3901586 | 27.0854134 | 13.86021 | 2.3750365 | 8.62513 | 2.32568227 |
| Collagen Complex - 2 | 2.89675793 | 2.080032 | 0.55182341 | 2.19948848 | 0.80001228 | 1.12501724 | 1.94502976 | 2.480038 | 1.180018 | 1.5500235 | 1.36422699 |
| Recombinant Human GABA Receptor Epsilon (gABRe) | 2.32742887 | 7.27094583 | 2.02242507 | 2.86362742 | 2.5 | 10.9663663 | 3.11688409 | 11.805137 | 1.51252333 | 1.68335917 | 2.55003917 |
| Recombinant Human Glutamate Receptor 3 (GRIA3) | 5.17597594 | 8.10429167 | 7.50830718 | 4.54792158 | 2.65306131 | 6.1219874 | 14.287427 | 7.03361165 | 2.79170917 | 5.0709125 | 1.61252458 |
| Human OMGP/OMG Recombinant Protein | 2.47956321 | 2.86671042 | 3.93687713 | 3.42147125 | 2.42749735 | 2.5 | 2.57006217 | 4.39388378 | 2.1250325 | 5.10424583 | 1.2458525 |
| Human Cytochrome P450 1B1 (CYP1B1) Protein | 1.47031841 | 1.93060702 | 2.08483765 | 2.36683488 | 2.03331173 | 3.2847601 | 1.54328209 | 2.05893166 | 2.18568897 | 2.08785653 | 2.80193646 |
| SmD3 | 9.15529145 | 1.9300295 | 2.21209226 | 1.50042627 | 0.96001472 | 1.56002388 | 4.07506226 | 2.095032 | 1.780027 | 1.6050245 | 3.6032917 |
| Cardiolipin | 3.25938594 | 1.4600225 | 4.15546932 | 2.71099742 | 2.08003182 | 5.37008202 | 5.500084 | 1.9300295 | 1.1350175 | 3.740057 | 2.71979235 |
| CYP21A2 | 108.203941 | 9.18014 | 1.15643008 | 10.2642794 | 1.00001534 | 1.80502762 | 69.2160562 | 13.565205 | 1.905029 | 1.225019 | 4.94586245 |
| Pancreatic and duodenal homeobox 1(PDX1) | 8.11433461 | 2.225034 | 0.94049911 | 9.95311142 | 1.23501892 | 1.90002907 | 7.2301104 | 4.110063 | 2.740042 | 2.6550405 | 2.08315326 |
| Recombinant Human Acetylcholine receptor | 3.50817358 | 5.82925417 | 7.14285596 | 3.28388211 | 2.08378105 | 3.11244999 | 3.50307617 | 5.47583118 | 6.3542625 | 2.79170917 | 1.75002667 |
| Threonyl-tRNA Synthetase(PI-7) | 2.69624605 | 1.3450205 | 3.27255331 | 1.57715261 | 1.22501877 | 1.41502167 | 2.77004234 | 2.7050415 | 1.0850165 | 1.4650225 | 1.2472933 |
| Recombinant Dipeptidyl Peptidase 6 (DPP6) | 3.4071295 | 3.59588833 | 2.36295683 | 2.33152355 | 3.49087049 | 4.45532106 | 1.73103173 | 8.86707149 | 1.90002917 | 2.66254083 | 2.13336583 |
| SP100 | 1.93686001 | 1.240019 | 0.8205374 | 1.07416883 | 0.70001076 | 1.14501755 | 1.12001717 | 1.4600225 | 0.965015 | 1.250019 | 1.09138185 |
| Human Involucrin (iNV) Protein | 2.16841953 | 1.49530997 | 1.70076211 | 1.65769912 | 1.73222343 | 1.97571379 | 2.04673387 | 2.02441117 | 1.78829108 | 1.9868722 | 1.20683319 |
| SRP54 | 0.81058023 | 2.275035 | 7.64875343 | 2.17391304 | 0.95501465 | 1.23501892 | 2.65504059 | 1.110017 | 1.0650165 | 2.61004 | 4.55175411 |
| Ganglioside GM1 | 2.18891829 | 1.75419333 | 1.27906974 | 1.33845006 | 3.46133281 | 1.64407632 | 3.25017156 | 0.96110293 | 3.95839375 | 0.85001292 | 0.97918167 |
| OJ (Isoleucyl tRNA Synthetase) | 4.87627812 | 1.430022 | 2.94865709 | 5.52429656 | 1.16001778 | 2.17003319 | 2.91004448 | 3.150048 | 0.9950155 | 2.3850365 | 5.30099781 |
| OGDC-E2 | 1.36518783 | 1.31002 | 0.87092147 | 2.03751066 | 0.74001137 | 1.08001656 | 1.44502213 | 1.105017 | 1.0600165 | 1.29002 | 1.33824168 |
| Insulin | 1.84726965 | 1.3250205 | 2.92706433 | 1.11679457 | 0.7750119 | 0.99001518 | 1.67502564 | 1.5400235 | 1.105017 | 1.6050245 | 1.66305787 |
| MDA5 | 1.11774748 | 1.125017 | 5.99088282 | 1.82011936 | 0.7550116 | 1.28001961 | 1.86502853 | 2.345036 | 1.0900165 | 2.370036 | 2.64616722 |
| Intrinsic factor | 1.61262817 | 1.2000185 | 4.98560543 | 2.60869563 | 1.05501617 | 1.07501648 | 1.74002663 | 2.560039 | 1.185018 | 2.4000365 | 2.3776529 |
| Zinc transporter 8 | 6.26279711 | 1.7300265 | 7.00575939 | 1.6794544 | 1.09501678 | 1.86002846 | 8.21012535 | 1.775027 | 1.29502 | 1.845028 | 2.81940236 |
| Alpha-Myosin | 3.14846442 | 1.1450175 | 1.52111334 | 3.55072459 | 1.02501572 | 1.2600193 | 3.82505844 | 1.4550225 | 1.235019 | 1.625025 | 1.45517537 |
| Myeloperoxidase | 2.07337883 | 1.29502 | 4.37380093 | 1.57289004 | 0.90001381 | 1.27001946 | 2.65004051 | 1.190018 | 1.125017 | 2.095032 | 2.61152028 |
| PM/SCl100 | 1.72354969 | 1.565024 | 9.74088327 | 1.67092925 | 0.92001411 | 1.34002052 | 1.75002678 | 1.7950275 | 1.185018 | 4.0250615 | 5.11909932 |
| Recombinant Inositol-1,4,5-Trisphosphate Receptor Type 3 (ITPR3) | 2.01180704 | 1.46668917 | 1.8646181 | 1.46574313 | 2.10526327 | 2.44979921 | 2.50854396 | 3.99358016 | 1.01251542 | 0.975015 | 0.92501417 |
| Th/To antigen | 2.06058039 | 1.125017 | 2.4424196 | 2.71099742 | 0.81501251 | 1.08501663 | 1.51502319 | 1.3350205 | 1.095017 | 1.5850245 | 3.01429199 |
| Glomerular Basement Membrane | 0.99829367 | 1.2150185 | 2.95585468 | 2.90707584 | 0.82001259 | 1.18001808 | 1.61502472 | 1.245019 | 0.8100125 | 1.705026 | 3.02728486 |

**Table S3. A panel of Neuronal Zoomer antigens used to measure plasma antibody levels via protein microarray in Mn-exposed workers and control groups.**

| Neuonal Zoomer Antigen | 3000/2675 | 3001/2849 | 3002 | 3003 | 3004/4755 | 3005/5608 | 3006/5965 | 3007/6013 | 3008/5997 | 3009/6069 | 3010/6166 |
|---|---|---|---|---|---|---|---|---|---|---|---|
| Group | Workers | Workers | Workers | Workers | Workers | Workers | Controls | Controls | Controls | Controls | Controls |
| Recombinant Human CRMP5 Protein | 3.14032617 | 4.6917375 | 4.92109589 | 3.21789566 | 1.44468312 | 13.792671 | 9.55229142 | 4.89426142 | 4.18339583 | 27.6045875 | 3.02921292 |
| Human SEPT5 / Septin 5 Protein | 13.0177032 | 33.217175 | 16.6694338 | 11.8831899 | 3.03168577 | 1.16967869 | 5.6151753 | 31.1480378 | 37.725575 | 3.67505625 | 4.20839583 |
| Recombinant Calcium Channel Alpha 1B Subunit (CACNa1B) | 4.93528483 | 25.7003917 | 23.1021594 | 4.81233721 | 4.70461944 | 6.90010052 | 4.90430766 | 6.06117921 | 10.404325 | 3.25838292 | 3.57922125 |
| Recombinant Endothelin Receptor A (ETRA) | 4.33015339 | 47.279875 | 10.1287364 | 6.21115513 | 3.27604831 | 7.40712843 | 6.02529217 | 13.549852 | 5.95009167 | 5.09174583 | 7.67511667 |
| AQP4 Recombinant | 4.13601243 | 11.1001708 | 6.80232545 | 4.07899626 | 1.93340499 | 12.2138548 | 9.47026941 | 6.28587748 | 3.48338667 | 2.87087708 | 1.59169083 |
| Glycine Receptor Alpha 1 Recombinant Protein | 7.81221475 | 2.8833775 | 1.56561478 | 1.3992887 | 4.45757165 | 3.76757051 | 28.482569 | 2.43391338 | 0.95418125 | 0.89168042 | 2.41670375 |
| Recombinant Human NARG2 Protein (NMDA) | 5.26907282 | 4.2959 | 5.71843836 | 2.98717663 | 4.31525373 | 5.10793159 | 2.15652772 | 6.02152724 | 3.66255583 | 1.78336042 | 1.80419417 |
| Dopamine Receptor D1 (DRD1) Protein | 7.48751041 | 2.45420417 | 4.44767289 | 2.5 | 1.93340499 | 5.18323415 | 59.9367952 | 2.5 | 3.77089083 | 2.89171083 | 1.82919458 |
| Caspr2 Recombinant Protein Antigen | 5.1271563 | 2.35003583 | 1.51162806 | 2.47519647 | 2.78195542 | 2.64558212 | 33.8294639 | 2.44335433 | 1.07084958 | 0.96668167 | 1.2583525 |
| Recombinant Human GABA Receptor Epsilon (gABRe) | 2.32742887 | 7.27094583 | 2.02242507 | 2.86362742 | 2.5 | 10.9663663 | 3.11688409 | 11.805137 | 1.51252333 | 1.68335917 | 2.55003917 |
| Recombinant Human GFAP Protein | 1.53496835 | 1.48335583 | 1.87707638 | 1.78491176 | 2.20730414 | 2.36194789 | 1.94292533 | 1.42749279 | 1.16251792 | 1.0416825 | 1.09168333 |
| Recombinant Human HOMER3 protein | 4.16439528 | 7.8167875 | 6.79401993 | 3.87495269 | 2.55370568 | 1.31777105 | 3.25529794 | 3.14010551 | 8.7168 | 2.87504375 | 1.95419667 |
| Human PLP1 / Myelin PLP Protein Recombinant | 3.80335976 | 6.57926667 | 9.05315484 | 3.58667136 | 5.91836838 | 6.48343268 | 4.71633714 | 9.61480567 | 4.62507083 | 12.4085208 | 2.733375 |
| Recombinant Dipeptidyl Peptidase 6 (DPP6) | 3.4071295 | 3.59588833 | 2.36295683 | 2.33152355 | 3.49087049 | 4.45532106 | 1.73103173 | 8.86707149 | 1.90002917 | 2.66254083 | 2.13336583 |
| Recombinant Human Glutamate Receptor 3 (GRIA3) | 5.17597594 | 8.10429167 | 7.50830718 | 4.54792158 | 2.65306131 | 6.1219874 | 14.287427 | 7.03361165 | 2.79170917 | 5.0709125 | 1.61252458 |
| Recombinant Human PNMA1 Protein | 2.98478588 | 5.970925 | 7.2799012 | 4.08086819 | 2.92159078 | 8.17520154 | 8.05878517 | 4.09365606 | 3.03337958 | 2.54587208 | 4.55840417 |
| Recombinant Human GRP78/HSPA5 Protein | 3.16303359 | 3.07504708 | 4.71760648 | 1.9412202 | 2.95918419 | 3.17520004 | 4.10970751 | 2.95128472 | 3.63338875 | 2.20003375 | 1.73336 |
| Human KCNA1 Protein | 3.34468609 | 5.30424583 | 6.88538066 | 4.35136619 | 3.49892622 | 7.55271104 | 5.59296101 | 7.33950205 | 5.7500875 | 2.41253708 | 1.74586 |
| Cluster of differentiation 11b (CD11b) | 2.46593945 | 1.86252833 | 1.66528227 | 1.54483308 | 2.05692808 | 2.86646479 | 2.22146352 | 2.61706961 | 1.25418583 | 0.93751417 | 1.10001667 |
| Recombinant GRB2 Related Adaptor Protein 2 (GRAP2) | 3.31743856 | 2.67087417 | 1.83139518 | 1.95993996 | 2.93233175 | 9.22439738 | 1.35680137 | 3.73489449 | 1.74586 | 1.23751875 | 1.07918333 |
| Recombinant Human AIF-1/Iba1 Protein | 3.30267834 | 3.95006042 | 5.20764051 | 3.87588865 | 2.65843179 | 3.36596269 | 4.01913984 | 2.65483339 | 3.55005417 | 3.06671333 | 1.27501958 |
| Recombinant Inositol-1,4,5-Trisphosphate Receptor Type 3 (ITPR3) | 2.01180704 | 1.46668917 | 1.8646181 | 1.46574313 | 2.10526327 | 2.44979921 | 2.50854396 | 3.99358016 | 1.01251542 | 0.975015 | 0.92501417 |
| Recombinant Human GAD2/GAD65 Protein | 3.12897303 | 3.14171458 | 1.8978406 | 3.25346279 | 4.59452176 | 5.01254901 | 2.5 | 4.78096819 | 1.30418667 | 0.97918167 | 1.61252458 |
| Recombinant Adaptor Related Protein Complex 3 Beta 2 (AP3b2) | 2.02202486 | 1.98336375 | 1.92691034 | 1.85885438 | 2.55907644 | 2.16616473 | 1.38926892 | 2.16389858 | 1.27918625 | 0.86668 | 1.08335 |
| Human MAG/Siglec-4a Protein | 4.03950847 | 3.32505083 | 15.7931889 | 2.89872703 | 2.22073036 | 1.24246999 | 1.92241983 | 4.94146614 | 2.70420792 | 5.20424583 | 1.65002542 |
| Recombinant Human Amphiphysin/AMPH Protein | 1.96071677 | 2.67504083 | 5.0830577 | 2.01656676 | 2.72287737 | 3.45883527 | 1.23376648 | 2.33761386 | 1.39168792 | 1.98753042 | 1.30835333 |
| Recombinant Human Dopamine D2R/DRD2 | 2.51930034 | 3.75422417 | 3.4260797 | 3.90256411 | 2.98066614 | 2.07831316 | 1.33629552 | 4.24848945 | 2.25003458 | 1.51252333 | 1.30418667 |
| Recombinant Human CDR2 Protein | 2.73274277 | 22.1253375 | 4.90863761 | 2.3947021 | 3.86949522 | 5.66766953 | 7.50854567 | 3.17975937 | 3.57088792 | 3.57922125 | 2.92921125 |
| Human EEF1A1 Recombinant Protein | 1.53269772 | 1.82086125 | 1.43272437 | 1.57759231 | 1.58700318 | 2.10090368 | 2.29323277 | 2.44524252 | 2.4625375 | 0.82084583 | 0.975015 |
| Recombinant Human PNMA2 Protein | 2.39554884 | 2.44170417 | 2.44186052 | 1.71284155 | 2.69065445 | 1.94779131 | 2.99555817 | 3.17787118 | 1.43335542 | 1.14168417 | 1.204185 |
| Human NOVA1 Protein | 1.8358311 | 1.8958625 | 2.41279078 | 2.36568673 | 2.39795913 | 3.04969994 | 1.85406815 | 1.36329323 | 1.51252333 | 1.80419417 | 1.67085875 |
| Recombinant Human Titin protein | 2.54541256 | 2.27503458 | 4.52242673 | 2.8818792 | 2.53759395 | 10.4216858 | 3.18352697 | 2.39614772 | 1.93752958 | 1.35002083 | 2.52920542 |
| Recombinant Human S100B | 3.73410447 | 66.226 | 4.64285679 | 6.88459106 | 2.8974209 | 2.85391478 | 3.46377396 | 3.06835433 | 3.48338667 | 2.97921208 | 1.63335833 |
| Recombinant Human SNCA | 1.62465929 | 1.00834875 | 1.66943504 | 1.07918364 | 1.32921603 | 0.97389553 | 3.01264608 | 1.31797613 | 1.43335542 | 0.80834583 | 0.86251333 |
| Recombinant Human CD68/SR-D1 Protein | 2.75544905 | 1.75419333 | 2.75747489 | 2.98951654 | 1.16272824 | 1.29769079 | 0.87662352 | 2.41691968 | 2.44170417 | 1.37085417 | 0.65834333 |
| Recombinant Human BACE-1 Protein | 3.58310544 | 4.045895 | 4.53903778 | 3.40228351 | 2.59667038 | 3.6244979 | 6.09022626 | 2.91163275 | 7.004275 | 3.44588583 | 1.73752667 |
| Human Tau-441 / 2N4R Protein | 6.41008041 | 5.02090833 | 4.99999834 | 4.70656853 | 2.68528396 | 4.6234937 | 3.7491456 | 4.37311181 | 4.62090417 | 4.2334 | 1.56669083 |
| Recombinant Human Enolase 2/Neuron-specific Enolase Ptotein | 4.02247876 | 4.4709 | 6.96428726 | 4.05934053 | 3.44253611 | 3.05973995 | 2.8827762 | 3.20052945 | 4.6667375 | 2.01253083 | 3.57922125 |
| Ganglioside GM1 | 2.18891829 | 1.75419333 | 1.27906974 | 1.33845006 | 3.46133281 | 1.64407632 | 3.25017156 | 0.96110293 | 3.95839375 | 0.85001292 | 0.97918167 |
| Recombinant Bone Morphogenetic Protein 1 (BMP1) | 3.09037236 | 1.9208625 | 1.85631216 | 0.52087229 | 3.47475903 | 2.02058237 | 1.48325401 | 1.46336876 | 1.47085583 | 0.92501417 | 1.29168625 |
| Recombinant Leucine-rich glioma-inactivated protein 1 (LGI1) | 2.70322347 | 2.1375325 | 1.76079743 | 1.18214163 | 1.78571444 | 1.75702816 | 1.28503109 | 2.03549795 | 1.00834875 | 0.88751375 | 0.84167958 |
| 5-Hydroxytryptamine (5-HT) | 2.35808348 | 5.91259167 | 5.36544956 | 4.24372838 | 3.66272877 | 5.8659647 | 3.34073924 | 1.83345946 | 2.10419875 | 1.62919167 | 2.65420708 |
| Recombinant Human MBP Protein | 3.54677539 | 3.29588375 | 5.2200988 | 3.23895534 | 1.37218047 | 3.29568264 | 3.40909091 | 3.24962236 | 1.96669667 | 2.52087167 | 1.08335 |
| rhCarbonic Anhydrase VIII | 3.24477733 | 2.7750425 | 4.65116231 | 2.98109286 | 3.61976488 | 3.81275055 | 3.09637859 | 2.65105701 | 2.77920917 | 2.74170875 | 1.10835042 |
| Recombinant Human Galectin 3 Protein | 3.46843759 | 3.83339208 | 5.0041511 | 2.94084591 | 2.51611173 | 4.7540163 | 2.75974148 | 4.06155685 | 4.89590833 | 2.28336833 | 1.6791925 |
| Recombinant Human Mannan Binding Lectin/MBL Protein | 4.1984547 | 3.28755042 | 5.46926858 | 3.88197289 | 3.2626221 | 3.12751 | 3.51162013 | 4.32590709 | 2.78337583 | 5.34591667 | 1.65835875 |
| POLY(L-GLUTAMIC ACID) | 1.33742031 | 1.65835875 | 1.59883728 | 1.02957705 | 1.726638 | 2.24146579 | 1.42686303 | 1.16125398 | 1.17918458 | 0.88751375 | 1.60835792 |
| Human GRM1 / MGLUR1 Protein | 1.65077151 | 1.71252625 | 1.47425239 | 1.78023194 | 3.40494001 | 2.67319214 | 1.81134665 | 2.03738614 | 1.33335375 | 0.92918083 | 1.14168417 |
| Tubulin Beta (TUBb) | 1.84831955 | 2.00003042 | 2.5 | 2.5290149 | 2.88130944 | 3.31325265 | 1.82843457 | 2.08081638 | 1.45835583 | 1.18335125 | 1.31668667 |
| Recombinant Human RAGE Protein | 2.10830986 | 1.42918833 | 1.71511623 | 1.33470621 | 1.11439305 | 1.566265 | 0.73479175 | 1.73904869 | 0.87084667 | 0.98334833 | 0.95001458 |
| alpha-2B Adrenergic R/ADRA2B Recombinant Protein Antigen | 1.46571306 | 1.204185 | 1.75664467 | 1.87710569 | 1.36143949 | 1.18975895 | 1.39952185 | 2.12424472 | 1.12918417 | 0.90834708 | 0.76251167 |
| Human Brain Cerebellum Protein | 1.25681188 | 1.36252083 | 2.03073101 | 1.17512142 | 2.33351247 | 1.45080316 | 1.35167499 | 1.11216031 | 0.89168042 | 0.975015 | 0.88751375 |
| GM2 | 1.74613902 | 1.23335208 | 1.85631216 | 1.95385619 | 1.80451141 | 1.76957842 | 1.29186642 | 1.15181285 | 0.770845 | 0.81667917 | 0.758345 |
| Human DBH/Dopamine beta-Hydroxylase Protein | 1.79609397 | 1.66669208 | 1.94352179 | 2.39563807 | 2.76047347 | 3.30321264 | 1.37388963 | 1.82401852 | 1.21251875 | 1.07501667 | 1.05001625 |
| Beta-Amyloid (25-35) | 1.75635799 | 2.42920375 | 2.16777418 | 3.01151218 | 1.97099894 | 2.16365473 | 1.08168179 | 1.44637487 | 1.38335458 | 1.19585167 | 0.92501417 |
| Human STK23/MSSK1/SRPK3/SPRY3 recombinant protein | 2.36262474 | 2.26253458 | 3.90780739 | 2.73633435 | 1.57626221 | 1.89759052 | 2.08304967 | 1.5143508 | 1.97503 | 1.95836333 | 1.2458525 |
| Recombinant Human Acetylcholine receptor | 3.50817358 | 5.82925417 | 7.14285596 | 3.28388211 | 2.08378105 | 3.11244999 | 3.50307617 | 5.47583118 | 6.3542625 | 2.79170917 | 1.75002667 |
| Recombinant Human Beta-amyloid 42 | 4.64463991 | 4.46673333 | 4.90033209 | 4.6710028 | 2.33619772 | 1.40311237 | 2.98530542 | 4.66389858 | 5.2209125 | 2.52920542 | 1.94169625 |
| Recombinant Human Recoverin Protein | 3.96003536 | 3.56255458 | 5.75996596 | 3.37981989 | 2.07572505 | 2.14859447 | 2.76828544 | 3.63481858 | 3.47088625 | 2.64587375 | 0.92918083 |
| Human Siglec-1 / CD169 Protein | 2.15485887 | 3.02921292 | 4.33554836 | 3.45797434 | 2.93233175 | 3.58433787 | 2.93062237 | 1.91465197 | 1.88752875 | 1.95003 | 1.21251875 |
| Human ELAVL3 / HUC Protein | 3.48319667 | 2.46670417 | 3.77491698 | 3.62457839 | 2.05692808 | 1.61646579 | 2.05912488 | 3.05136063 | 2.52087167 | 1.61252458 | 1.44168875 |
| Recombinant Human NRXN3 Protein | 2.5 | 3.32088417 | 4.70930095 | 4.18008185 | 2.75778823 | 2.93674735 | 2.90840808 | 3.57439653 | 1.96253 | 3.27921667 | 1.408355 |
| Human OMGP/OMG Recombinant Protein | 2.47956321 | 2.86671042 | 3.93687713 | 3.42147125 | 2.42749735 | 2.5 | 2.57006217 | 4.39388378 | 2.1250325 | 5.10424583 | 1.2458525 |
| Recombinant Human Neurofascin Protein | 2.36603068 | 3.12088083 | 3.97009963 | 3.08451883 | 3.12298674 | 2.30421684 | 3.20403418 | 4.32968346 | 2.60837333 | 3.46671958 | 2.33336917 |

**Table S4. The 5 top enriched metabolite sets for ANCOVA results in metabolite enrichment analysis using RaMP-DB in plasma samples of Mn-exposed workers compared than Control group.** Hits: The metabolites in the set that are significantly changed in the experimental data. P < 0.05, * FDR < 0.05.

| Enriched Metabolite Sets | Hits |
|---|---|
| **Biogenic amine synthesis*** | **Choline, Glutamic acid, Phenylalanine** |
| **Transmission across Chemical Synapses*** | **Choline, Homovanillic acid, 3,4-Dihydroxybenzeneacetic acid, Glutamic acid** |
| **Neuronal System*** | **Choline, Homovanillic acid, 3,4-Dihydroxybenzeneacetic acid, Glutamic acid** |
| **22q11.2 copy number variation syndrome*** | **Homovanillic acid, 3,4-Dihydroxybenzeneacetic acid, Glutamic acid** |
| **Neurotransmitter clearance*** | **Choline, Homovanillic acid, 3,4-Dihydroxybenzeneacetic acid** |

**Table S5. The 5 top enriched metabolite sets for linear regression results in metabolite enrichment analysis using RaMP-DB in plasma samples of Mn-exposed workers compared than Control group**. Hits: The metabolites in the set that are significantly changed in the experimental data. P < 0.05.

| Enriched Metabolite Set | Hits |
|---|---|
| **SLC-mediated transmembrane transport** | L-Carnitine, Citric acid, Hypoxanthine, L-Tyrosine, Proline, Lactic acid, Uracil, Creatinine, N-Acetylneuraminic acid, L-Aspartic acid, L-Valine, L-Threonine, 4-Hydroxyproline |
| **Transport of small molecules** | L-Carnitine, Citric acid, Hypoxanthine, L-Tyrosine, Proline, Lactic acid, Uracil, Creatinine, N-Acetylneuraminic acid, L-Aspartic acid, L-Valine, L-Threonine, 4-Hydroxyproline |
| **Biochemical pathways: part I** | Pipecolic acid, Citric acid, Hypoxanthine, L-Tyrosine; Proline, L-Threonine, Lactic acid, L-Aspartic acid, N6-Acetyl-L-lysine, Xanthine, Uracil, 4-Hydroxyproline, L-Valine; Citrulline; 5-Aminolevulinic acid, Methionine sulfoxide, Carglumic acid |
| **Transport of bile salts and organic acids, metal ions and amine compounds** | L-Carnitine, Citric acid, L-Tyrosine, Proline, Lactic acid, Creatinine, L-Valine, L-Threonine, 4-Hydroxyproline |
| **Transport of inorganic cations/anions and amino acids/oligopeptides** | L-Tyrosine, Proline, Lactic acid, N-Acetylneuraminic acid, L-Aspartic acid, L-Valine, L-Threonine, 4-Hydroxyproline |

**Table S6. Fold changes of significantly distinct antibodies.** Fold changes in the antibodies (expressed in signal intensity unit or arbitrary units (AU)) targeting two panels of AI (Autoimmune) and NZ (neuronal Zoomer) proteins with significantly different abundance in the serum samples of Mn exposed workers identified using protein microarray. ** ($p < 0.01$) * ($p < 0.05$).

| Antigen Name | Fold Change |
| --- | --- |
| 5-Hydroxytryptamine (5-HT) (Serotonin) | 1.97* |
| Beta-Amyloid (25-35) | 1.86** |
| Recombinant Human Amphiphysin/AMPH Protein | 1.81* |
| GM2 | 1.80** |
| Human DBH/Dopamine Beta-Hydroxylase Protein | 1.77* |
| Human Siglec-1 / CD169 Protein | 1.64* |
| Tubulin Beta (TUBb) | 1.60* |
| Recombinant Adaptor Related Protein Complex 3 Beta 2 (AP3b2) | 1.54* |
| Human Brain Cerebellum Protein | 1.53* |
| rhCarbonic Anhydrase VIII | 1.42* |
| Human NOVA1 Protein | 1.42* |
| Recombinant Human GFAP Protein | 1.41* |
| Anti Mitochondria antibodies M2 | 0.74* |
